# COVID-19 infection and vaccination rates in healthcare workers in British Columbia, Canada: A Longitudinal Urban versus Rural Analysis of the Impact of the Vaccine Mandate

**DOI:** 10.1101/2022.01.13.22269078

**Authors:** Annalee Yassi, Stephen Barker, Karen Lockhart, Deanne Taylor, Devin Harris, Harsh Hundal, Jennifer M. Grant, Arnold Ikedichi Okpani, Sue Pollock, Stacy Sprague, Chad Kim Sing

**Author notes:** Corresponding author: Dr. Annalee Yassi, 430-2206 East Mall, UBC, Vancouver, BC, V6T 1Z3. Disclosures: AY, DT, DH, HH, JMG, AIO, SP, SS, CKS are all employees and/or contractors of VCH and/or IH.

## Abstract

**Purpose:** Healthcare workers (HCWs) play a critical role in responding to the COVID-19 pandemic. Early in the pandemic, urban centres were hit hardest globally; rural areas gradually became more impacted. We compared COVID-19 infection and vaccine uptake in HCWs living in urban versus rural locations within, and between, two health authorities in British Columbia (BC), Canada. We also analyzed the impact of a vaccine mandate for HCWs.

**Methods:** We tracked laboratory-confirmed SARS-CoV-2 infections, positivity rates, and vaccine uptake in 29,021 HCWs in Interior Health (IH) and 24,634 HCWs in Vancouver Coastal Health (VCH), by occupation, age, and home location, comparing to the general population in that region. We then evaluated the impact of infection rates as well as the mandate on vaccination uptake.

**Results:** By October 27, 2021, the date that unvaccinated HCWs were prohibited from providing healthcare, only 1.6% in VCH yet 6.5% in IH remained unvaccinated. Rural workers in both areas had significantly higher unvaccinated rates compared with urban dwellers. Over 1,800 workers, comprising 6.4% of rural HCWs and 3.3% of urban HCWs, remained unvaccinated and set to be terminated from their employment. While the mandate prompted a significant increase in second doses, the impact on the unvaccinated was less clear.

**Conclusions:** As rural areas often suffer from under-staffing, loss of HCWs could have serious impacts on healthcare provision as well as on the livelihoods of unvaccinated HCWs. Greater efforts are needed to understand how to better address the drivers of rural-related vaccine hesitancy as the pandemic continues.

## Introduction

Healthcare workers (HCWs) are on the frontlines of the world’s fight against COVID-19, striving to care for COVID-19 patients while also trying to manage regular and ongoing healthcare demands during a pandemic. Significant pressures faced by HCWs during the COVID-19 pandemic include an increased health system burden, risk of infection, burnout, mental health stresses, risk of healthcare worker shortages, and concerns about family transmission^1^. HCWs in rural settings face even greater pressures, as there are often even greater staffing shortages^2^. British Columbia (BC), Canada, instituted mandatory vaccination of healthcare workers; long-term care workers were to be vaccinated before October 12, 2021^3^, and those working in acute care and other publicly-funded healthcare facilities were to be vaccinated by October 26, 2021^4^. Vaccine mandates have been discussed for decades for healthcare workers for other communicable diseases such as influenza^5-7^ and, while some jurisdictions have chosen to allow those working in healthcare to remain unvaccinated against COVID-19, the upswing in cases across the world and the new Omicron variant^8^, has many countries moving towards mandating vaccination for HCWs during this pandemic ^9-11^. As vaccine uptake is now well-established to be an important determinant of COVID-19 case rates and morbidity, we sought to understand the extent to which rurality impacted COVID-19 rates, vaccine uptake and drivers of vaccination within two of the five large health authorities located in British Columbia (BC), Canada – one mainly rural, located in the interior of the province, namely Interior Health (IH), and one more urban, namely Vancouver Coastal Health (VCH). Moreover, as BC brought in a mandate that required vaccination of all HCWs, we sought to compare and contrast rural and urban differences not only in 1) COVID-19 rates and 2) vaccine uptake, within and between health regions and whether there were 3) any differences within occupational group, and 4) by age-group, but also 5) impact of higher COVID rates in the previous month on subsequent vaccine uptake; and finally, 6) the impact of the mandated vaccination (on threat of termination of employment) on vaccine uptake in both settings.

HCWs from rural areas reported significantly less willingness to take a vaccine in the early phases of the pandemic (26%), compared to their suburban (35%) and urban (37%) peers^12^, with this trend persisting in the general U.S. population throughout the pandemic^13^. Murthy and colleagues found adult COVID-19 vaccination coverage lower in rural (38.9%) than in urban counties (45.7%) overall and among adults aged 18–64 years (29.1% rural, 37.7% urban), those aged ≥65 years (67.6% rural, 76.1% urban), women (41.7% rural, 48.4% urban), and men (35.3% rural, 41.9% urban)^14^. Access and acceptance disparities have been documented – i.e. people from rural locations having to travel outside their counties to receive a vaccine^15^. This trend has been seen elsewhere as well^16-18^. In a study to assesses the associations of age, gender, and level of education with vaccine acceptance, Lazarus et al. used a random sample of 13,426 participants selected from 19 high-COVID-19 burden countries in June 2020. Based on univariable and multivariable logistic regression, several noteworthy trends emerged: women in France, Germany, Russia, and Sweden were significantly more likely to accept a vaccine than men in these countries. Older (≥50) people in Canada, Poland, France, Germany, Sweden, and the UK were significantly more favorably disposed to vaccination than younger respondents, but the reverse trend held in China. Highly educated individuals in Ecuador, France, Germany, India, and the US reported that they will accept a vaccine, but higher education levels were associated with lower vaccination acceptance in Canada, Spain, and the UK^19^.

Data on barriers and facilitators to uptake of COVID-19 vaccines within Canada is scarce as Canada has had strong vaccine uptake (76.3% of country as of December 13, 2021^20^) however the range is 61.2% of those in Nunavut to 84.9% in Newfoundland and Labrador^21^. In a recent systematic review related to vaccine uptake in children aged up to 7 years old, focused within a Canadian context, the authors found that between 50% and 70% of children are completely vaccinated at 2 years old, with up to 97% having received at least one vaccine, and 2–5% receiving no vaccines. This review found that trust and access to health care providers is significantly associated with vaccine uptake, likely more important than parents’ vaccine knowledge, and may compensate for challenges related to socio-economic status and family dynamics^22^.

Globally, there are disparities noted in uptake of childhood vaccines with those living in rural locations being less likely to vaccinate their children^23-25^. Rurality itself is defined as an important social determinant of health^26^. As such, there is a particular need to assess the impact not only of COVID-19 infections, but also of how COVID-19 vaccination policies are working in rural compared to urban areas in one of the first jurisdictions to implement a vaccine mandate specifically for healthcare workers.

## Methods

### Cohort description

The cohort included all healthcare workers employed by Interior Health (n=29,021) and Vancouver Coastal Health (n= 24,634) for at least one day between March 1, 2020 and November 11, 2021. When analysis considered a specific date within that interval, a subset of the cohort was used, excluding those that did not have an active appointment on that date.

### Database

Healthcare worker records were obtained from the provincial Workplace Health Indicator Tracking and Evaluation (WHITE™) database. Following ethics approval (UBC Behavioural Ethics Certificate H21-01380) the data fields extracted included worker demographics (age group, gender, home location), job details (job title, job category, subsector, job location, job start date, and if applicable, job end date), severe acute respiratory syndrome coronavirus 2 (SARS-CoV-2) PCR testing information (date, test result) and COVID-19 vaccination status (date of vaccine and type of vaccine). Data on the background communities were obtained from the B.C. Centre for Disease Control, and included vaccinations (daily vaccination dose totals by health region) and infection totals (daily positive and negative test counts by health region, including age group for positive cases), with regional population data obtained from Statistics Canada. Home and work locations were provided as the local health area (LHA), a subdivision of the regional health authorities; there were further classified as either urban or rural based on population size of the LHA, summarised in Table S1. Jobs were classified into six categories: nurses, licensed practical nurses (LPNs)/ care aides, administration, allied health, support staff, others/unknown. Ages were classified into four categories: 39 and under, 40-49, 50-59 and 60 and over.

### Statistical analysis

For each health authority, we calculated the SARS-CoV-2 infection rate (per 100,000 population) over time as a 7-day moving average, also plotting the cumulative proportion with 2 or more doses of vaccine, for both HCWs and the background community from March 1, 2020 to November 11, 2021. The background community infection rates were both region and age-adjusted by weighting positive cases to match the residence and age-range distribution of the workforce (see supplementary table S2). Over the same period, we plotted the same variables for HCWs alone, comparing those residing in rural locations with those residing in urban locations.

SARS-CoV-2 infection rates and COVID-19 vaccination status were tabulated by health authority, occupation group, home residence type (urban/rural) and age group. To address our first four key research questions (COVID infection and vaccine uptake respectively, and any differences in this regard between occupational or age groups), effect size models using logistic regression were used to calculate odds ratios. The dependent variable was whether the individual had received at least one dose of vaccine prior to a specified date or not, or whether the individual had tested positive for SARS-CoV-2 at least once prior to a specified date. The variables of interest included the home residence type (rural or urban), occupation group and age group. These values were calculated on the day before the vaccine mandate announcement, September 12, 2021, and the day the mandate took effect on October 27, 2021.

To ascertain the extent to which COVID-19 rates in the period prior to vaccination drove vaccination rates (question 4), we considered the period when vaccination was available to healthcare workers, from December 15, 2020 to November 11, 2021. For each date in the observation period, we counted one observation per HCW, where the response was 0 if the HCW was unvaccinated on that date, 1 if they received the first dose on that date, excluding all days after the first dose. The variable of interest was the community infection rate for the home region of the HCW on that date; for this, we calculated the daily 14 day moving average background community SARS-CoV-2 infection rate for each region. To account for repeated measures on a single HCW, conditional logistic regression was used, with each individual HCW making up one of the strata. Anyone who had tested positive prior to December 15, 2020 was also excluded from the calculation.

To examine the final question, the extent to which the mandate for compulsory vaccination of all HCWs drove vaccination uptake, we examined the period from July 1, 2021 to October 27, 2021, using segmented regression analysis^27^ of the interrupted time series (ITS) to estimate the immediate and sustained effects on the rate of vaccination following the announcement, where the rate is measured as the proportion of workers that received the dose on a given day out of the total number of workers that had not yet received that dose. For workers in the long-term care sector, the mandate took affect a few days earlier; therefore, long-term care workers were excluded from this analysis.

## Results

To answer both question #1 and #2 regarding COVID infection and vaccination rates in HCWs, Figure 1 shows the HCW and background community SARS-CoV-2 infection rates in Interior Health displayed against vaccination status in these respective groups. The initial small peak shown in Figure 1 could be related to a combination of increased case finding activities in HCWs as well as the less clear guidance on personal protective equipment (PPE) use and less availability of PPE than was the case subsequently. In September and October 2020, HCW infections trailed off significantly, even more so than community infections. In the second wave (beginning towards the end of October 2020), again, we see a peak wherein HCW COVID-19 rates exceeded community rates, again possibly related to increased case finding associated with the policy of testing asymptomatic HCWs during outbreaks. Additionally, Figure 1 shows that HCW vaccination has been steadily higher than that of the general population.

**Figure 1:**
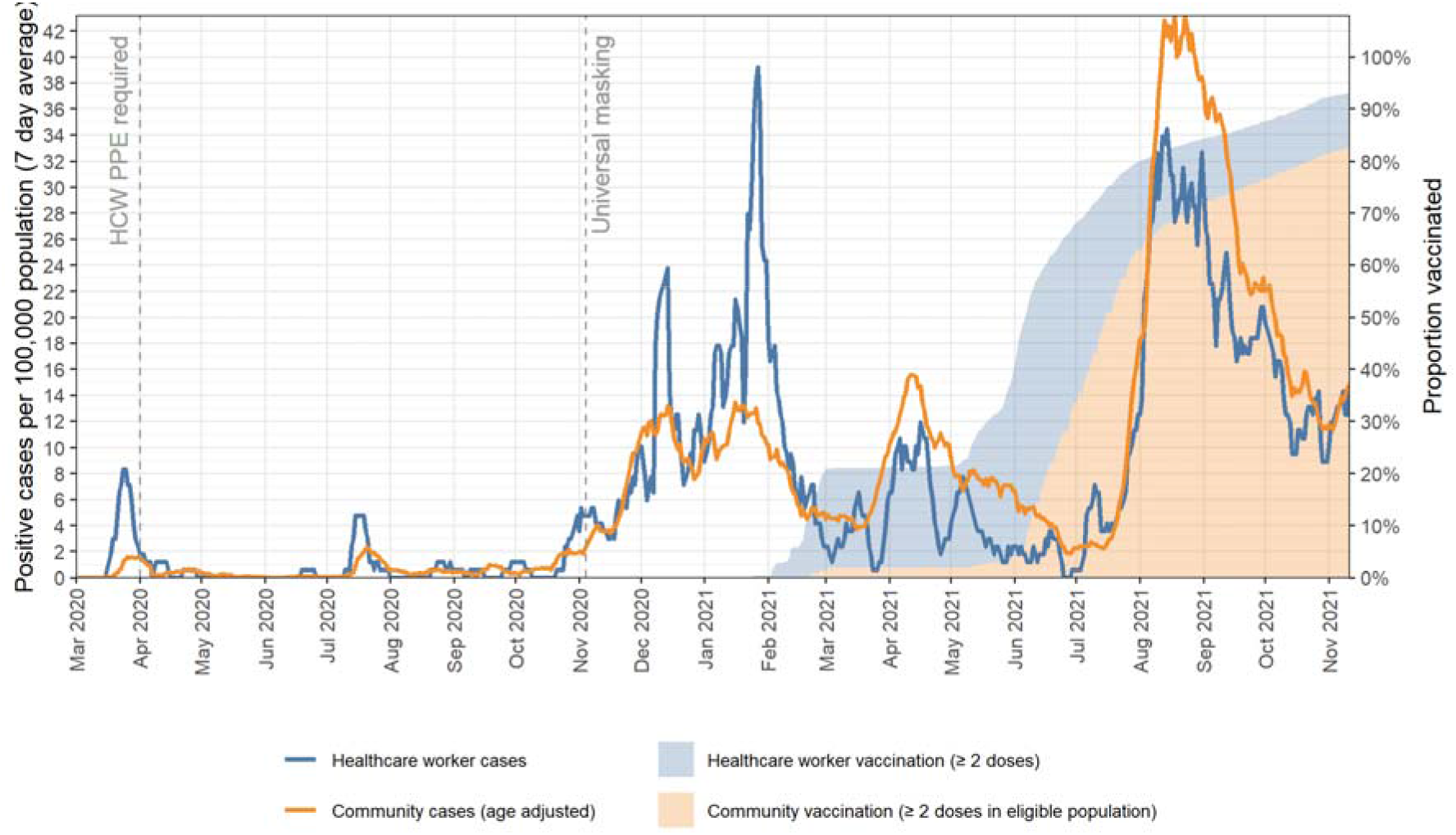
**Interior Health (IH)** COVID-19 case rate in healthcare workers and the age-adjusted community rate, showing the proportion fully vaccinated.

Figure 2 shows that while VCH experienced a larger initial impact, it did not experience the same intensity of infections in the fourth wave as IH (Figure 1), and that, unlike IH, the HCWs in VCH were largely protected in the third wave. The data suggest that this greater protection relates to the increased vaccination uptake compared to the community at this point in the pandemic. Studies reported elsewhere^28,29^, providing positivity rates, are consistent with the peaks in HCW infections in the early stages being related to increased testing, as the HCW positivity rates closely follow community rates in the first two waves.

**Figure 2:**
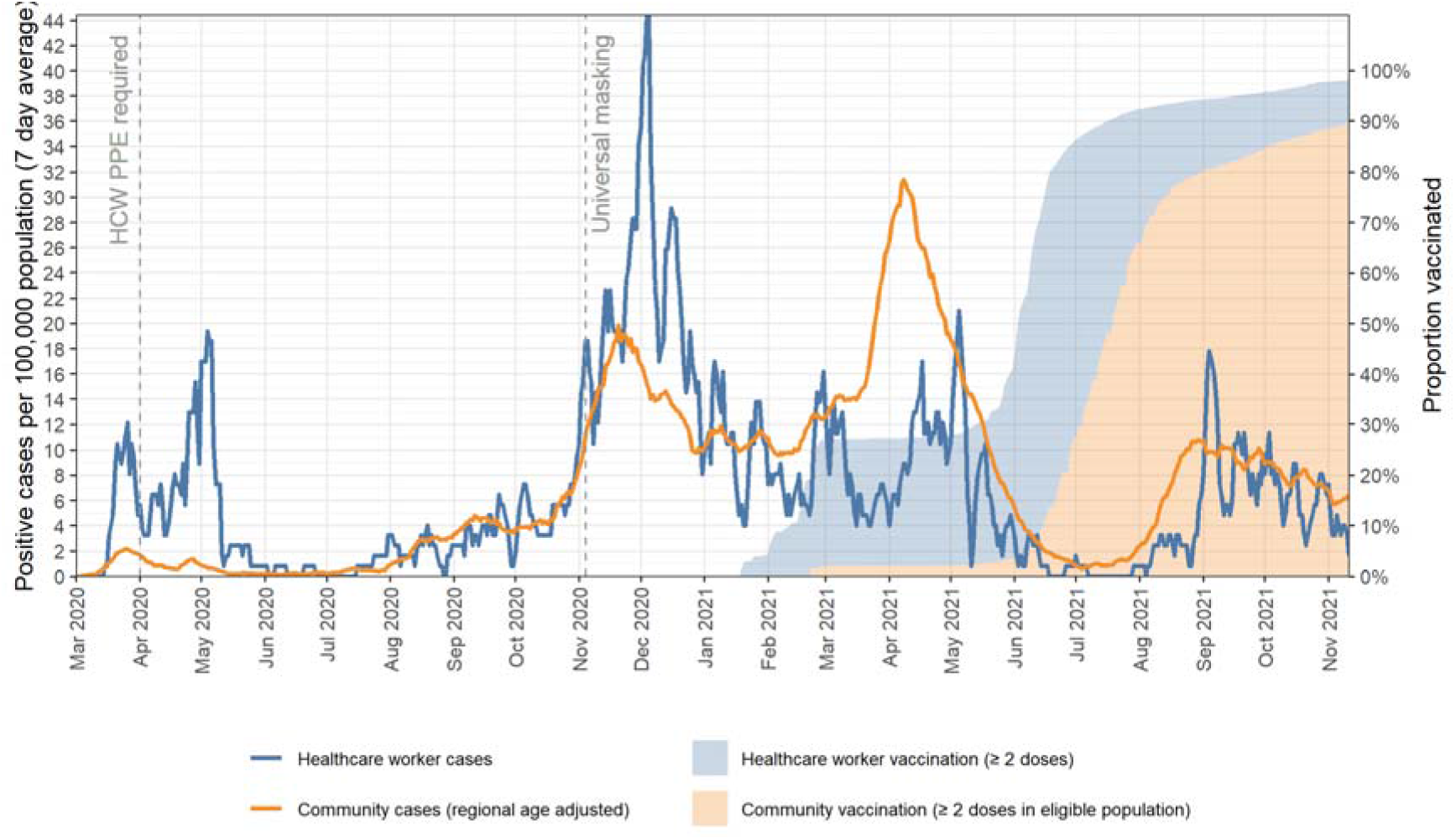
**Vancouver Coastal Health (VCH)** COVID-19 case rate in healthcare workers and the age adjusted community rate, showing the proportion fully vaccinated.

Regarding the urban-rural divide, Figure 3 shows the urban and rural breakdown of vaccination and cases in healthcare workers in the two jurisdictions (IH and VCH combined). Rural cases have followed a similar trend to their urban counterparts with the exception of spikes in the last two months (Sept. – Oct. 2021) where rural cases outpaced those in urban locations. Table 1 provides the rate of unvaccinated HCWs, comparing the two health regions, both separately and combined, divided by residence type (urban or rural), occupation and age group; Table 2 shows the same breakdown for SARS-CoV-2 infection rates.

**Figure 3:**
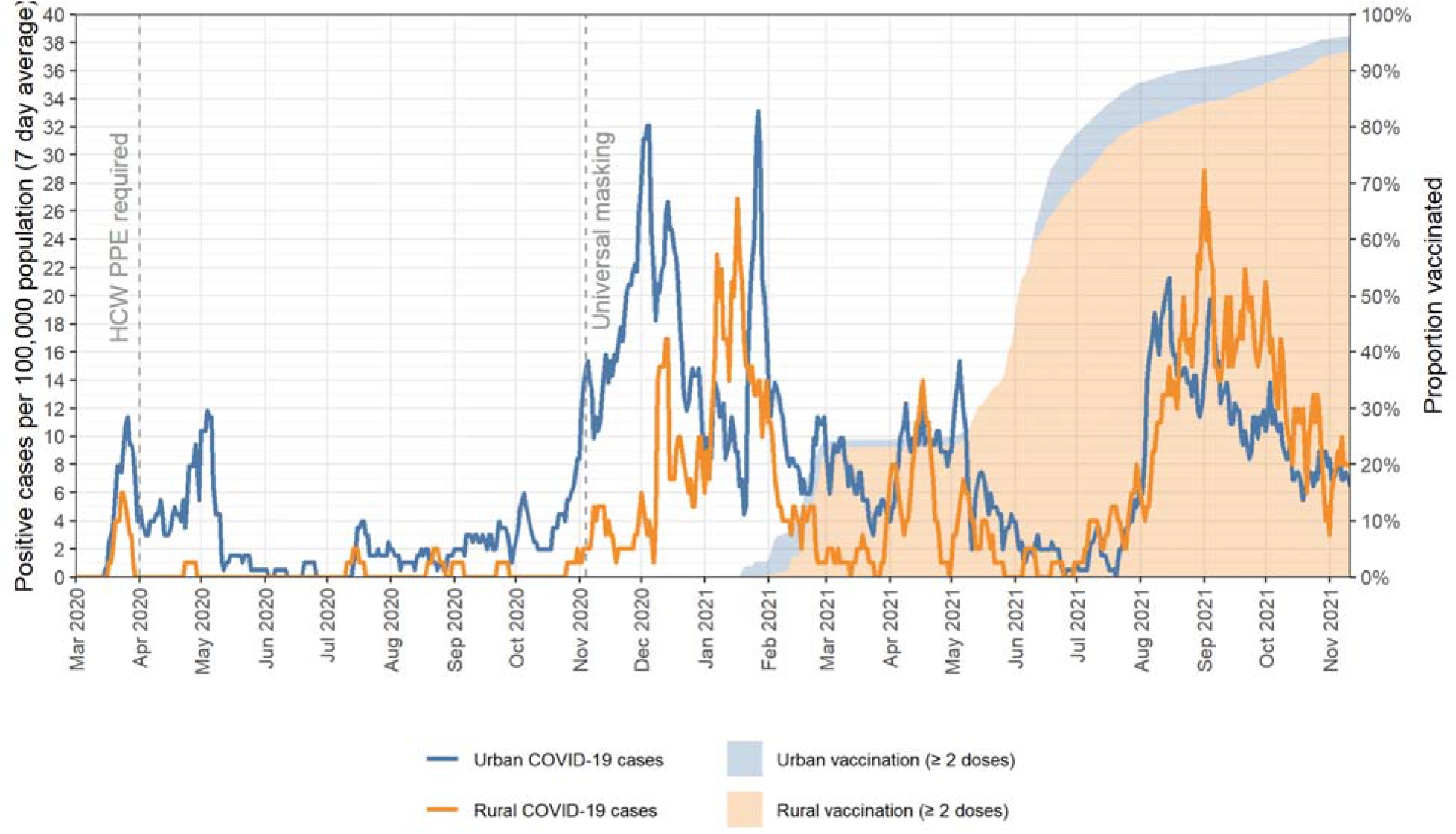
COVID-19 case rate and proportion vaccinated **comparing urban and rural populations**.

**Table 1:** Healthcare workers’ unvaccinated rate by region on the date of the announcement of the mandate and when it took effect, broken down by residence type, occupation and age group.

| Worksite region | Exposure Group | Remaining unvaccinated on October 27, 2021 |  |  | Remaining unvaccinated on September 12, 2021 |  |  |
| --- | --- | --- | --- | --- | --- | --- | --- |
|  |  | % | (#/n) | OR (95% CI) | % | (#/n) | OR (95% CI) |
| Interior Health (IH) | Urban | 6.1% | (695/11,366) | 0.90 (0.81, 1.00)* | 11.1% | (1,260/11,317) | 0.82 (0.76, 0.89)* |
|  | Rural | 6.8% | (862/12,752) | 1.11 (1.00, 1.23)* | 13.2% | (1,689/12,762) | 1.22 (1.13, 1.32)* |
|  | LPN / Care Aides | 7.6% | (567/7,470) | 1.30 (1.17, 1.45)* | 15.1% | (1,136/7,507) | 1.45 (1.34, 1.57)* |
|  | Nurses | 4.5% | (288/6,470) | 0.60 (0.53, 0.69)* | 7.6% | (493/6,479) | 0.51 (0.46, 0.56)* |
|  | Administration | 7.9% | (282/3,574) | 1.29 (1.13, 1.48)* | 13.9% | (492/3,543) | 1.19 (1.07, 1.32)* |
|  | Allied Health | 3.9% | (122/3,121) | 0.55 (0.46, 0.67)* | 6.9% | (214/3,117) | 0.49 (0.43, 0.57)* |
|  | Support | 8.2% | (262/3,203) | 1.35 (1.18, 1.55)* | 17.8% | (566/3,176) | 1.69 (1.52, 1.86)* |
|  | 39 and under | 6.8% | (721/10,563) | 1.11 (1.01, 1.24)* | 13.5% | (1,413/10,450) | 1.23 (1.14, 1.33)* |
|  | 40-49 | 6.5% | (360/5,508) | 1.02 (0.90, 1.15) | 11.7% | (643/5,476) | 0.94 (0.86, 1.03) |
|  | 50-59 | 6.0% | (320/5,353) | 0.90 (0.79, 1.02) | 10.8% | (580/5,385) | 0.83 (0.76, 0.92)* |
|  | 60 and over | 5.8% | (156/2,694) | 0.88 (0.74, 1.04) | 11.3% | (313/2,768) | 0.90 (0.80, 1.02) |
|  | Overall | 6.5% | (1,557/24,118) | - | 12.2% | (2,949/24,079) | - |
| Vancouver Coastal Health (VCH) | Urban | 1.4% | (232/16,736) | 0.35 (0.26, 0.45)* | 3.2% | (569/17,696) | 0.44 (0.37, 0.54)* |
|  | Rural | 3.9% | (70/1,795) | 2.89 (2.20, 3.79)* | 7.0% | (132/1,898) | 2.25 (1.85, 2.74)* |
|  | LPN / Care Aides | 1.9% | (84/4,383) | 1.25 (0.97, 1.61) | 4.1% | (189/4,559) | 1.23 (1.03, 1.45)* |
|  | Nurses | 1.3% | (83/6,318) | 0.73 (0.57, 0.94)* | 2.8% | (187/6,733) | 0.69 (0.58, 0.81)* |
|  | Administration | 1.8% | (66/3,605) | 1.16 (0.88, 1.53) | 4.2% | (157/3,703) | 1.25 (1.04, 1.50)* |
|  | Allied Health | 1.3% | (40/3,168) | 0.74 (0.53, 1.03) | 3.0% | (97/3,283) | 0.79 (0.64, 0.98)* |
|  | Support | 3.4% | (23/670) | 2.24 (1.45, 3.45)* | 8.0% | (55/690) | 2.45 (1.84, 3.26)* |
|  | 39 and under | 1.4% | (109/7,672) | 0.80 (0.63, 1.01) | 3.4% | (281/8,312) | 0.90 (0.78, 1.06) |
|  | 40-49 | 1.3% | (58/4,346) | 0.77 (0.58, 1.03) | 3.1% | (141/4,478) | 0.85 (0.70, 1.02) |
|  | 50-59 | 1.9% | (80/4,280) | 1.20 (0.93, 1.56) | 4.2% | (186/4,402) | 1.26 (1.06, 1.49)* |
|  | 60 and over | 2.5% | (55/2,233) | 1.64 (1.22, 2.21)* | 3.9% | (93/2,402) | 1.10 (0.88, 1.37) |
|  | Overall | 1.6% | (302/18,531) | - | 3.6% | (701/19,594) | - |
| Overall (Both VCH and IH combined) | Urban | 3.3% | (927/28,102) | 0.50 (0.45, 0.55)* | 6.3% | (1,829/29,013) | 0.47 (0.44, 0.51)* |
|  | Rural | 6.4% | (932/14,547) | 2.01 (1.83, 2.20)* | 12.4% | (1,821/14,660) | 2.11 (1.97, 2.26)* |
|  | LPN / Care Aides | 5.5% | (651/11,853) | 1.42 (1.29, 1.57)* | 11.0% | (1,325/12,066) | 1.55 (1.45, 1.67)* |
|  | Nurses | 2.9% | (371/12,788) | 0.57 (0.51, 0.64)* | 5.1% | (680/13,212) | 0.50 (0.46, 0.55)* |
|  | Administration | 4.8% | (348/7,179) | 1.14 (1.02, 1.29)* | 9.0% | (649/7,246) | 1.10 (1.00, 1.20)* |
|  | Allied Health | 2.6% | (162/6,289) | 0.54 (0.46, 0.64)* | 4.9% | (311/6,400) | 0.52 (0.46, 0.58)* |
|  | Support | 7.4% | (285/3,873) | 1.88 (1.65, 2.14)* | 16.1% | (621/3,866) | 2.32 (2.12, 2.55)* |
|  | 39 and under | 4.6% | (830/18,235) | 1.08 (0.99, 1.19) | 9.0% | (1,694/18,762) | 1.16 (1.09, 1.25)* |
|  | 40-49 | 4.2% | (418/9,854) | 0.96 (0.86, 1.08) | 7.9% | (784/9,954) | 0.92 (0.85, 1.00)* |
|  | 50-59 | 4.2% | (400/9,633) | 0.94 (0.84, 1.05) | 7.8% | (766/9,787) | 0.91 (0.84, 0.99)* |
|  | 60 and over | 4.3% | (211/4,927) | 0.98 (0.85, 1.13) | 7.9% | (406/5,170) | 0.93 (0.83, 1.03) |
|  | Overall | 4.4% | (1,859/42,649) | - | 8.4% | (3,650/43,673) | - |

\* = unadjusted odds ratio (OR) is significant at 95% confidence

**Table 2:** Healthcare workers’ SARS-CoV-2 cumulative infection rate by region on the date of the announcement of the mandate and when it took effect, broken down by residence type, occupation and age group.

| Worksite region | Exposure Group | Infected by October 27, 2021 |  |  | Infected by September 12, 2021 |  |  |
| --- | --- | --- | --- | --- | --- | --- | --- |
|  |  | % | (#/n) | OR (95% CI) | % | (#/n) | OR (95% CI) |
| Interior Health (IH) | Urban | 4.8% | (551/11,366) | 1.46 (1.28, 1.66)* | 4.3% | (482/11,317) | 1.64 (1.42, 1.88)* |
|  | Rural | 3.4% | (431/12,752) | 0.69 (0.60, 0.78)* | 2.6% | (338/12,762) | 0.61 (0.53, 0.70)* |
|  | LPN / Care Aides | 5.8% | (433/7,470) | 1.80 (1.59, 2.05)* | 4.9% | (367/7,507) | 1.83 (1.59, 2.10)* |
|  | Nurses | 3.6% | (233/6,470) | 0.84 (0.73, 0.98)* | 2.9% | (186/6,479) | 0.79 (0.67, 0.93)* |
|  | Administration | 2.6% | (92/3,574) | 0.58 (0.47, 0.73)* | 2.4% | (84/3,543) | 0.65 (0.52, 0.82)* |
|  | Allied Health | 2.8% | (86/3,121) | 0.64 (0.51, 0.80)* | 2.2% | (70/3,117) | 0.62 (0.48, 0.79)* |
|  | Support | 4.0% | (129/3,203) | 0.99 (0.82, 1.19) | 3.3% | (106/3,176) | 0.98 (0.79, 1.20) |
|  | 39 and under | 5.0% | (528/10,563) | 1.52 (1.34, 1.73)* | 4.2% | (444/10,450) | 1.56 (1.36, 1.80)* |
|  | 40-49 | 3.7% | (202/5,508) | 0.87 (0.74, 1.02) | 2.9% | (157/5,476) | 0.80 (0.67, 0.95)* |
|  | 50-59 | 3.7% | (197/5,353) | 0.88 (0.75, 1.03) | 3.1% | (169/5,385) | 0.90 (0.76, 1.07) |
|  | 60 and over | 2.0% | (55/2,694) | 0.46 (0.35, 0.61)* | 1.8% | (50/2,768) | 0.49 (0.37, 0.66)* |
|  | Overall | 4.1% | (982/24,118) | - | 3.4% | (820/24,079) | - |
| Vancouver Coastal Health (VCH) | Urban | 4.4% | (739/16,736) | 2.08 (1.50, 2.88)* | 4.1% | (725/17,696) | 2.34 (1.66, 3.31)* |
|  | Rural | 2.2% | (39/1,795) | 0.48 (0.35, 0.67)* | 1.8% | (34/1,898) | 0.43 (0.30, 0.60)* |
|  | LPN / Care Aides | 5.7% | (251/4,383) | 1.57 (1.35, 1.83)* | 5.3% | (242/4,559) | 1.57 (1.35, 1.84)* |
|  | Nurses | 4.0% | (250/6,318) | 0.91 (0.78, 1.06) | 3.6% | (241/6,733) | 0.88 (0.76, 1.03) |
|  | Administration | 3.9% | (140/3,605) | 0.90 (0.75, 1.09) | 3.6% | (132/3,703) | 0.90 (0.74, 1.09) |
|  | Allied Health | 2.9% | (93/3,168) | 0.65 (0.52, 0.81)* | 2.8% | (92/3,283) | 0.68 (0.54, 0.84)* |
|  | Support | 4.3% | (29/670) | 1.03 (0.71, 1.51) | 4.1% | (28/690) | 1.05 (0.72, 1.55) |
|  | 39 and under | 5.0% | (381/7,672) | 1.38 (1.19, 1.59)* | 4.7% | (388/8,312) | 1.44 (1.25, 1.67)* |
|  | 40-49 | 4.5% | (197/4,346) | 1.11 (0.94, 1.31) | 3.9% | (174/4,478) | 1.00 (0.84, 1.19) |
|  | 50-59 | 3.2% | (138/4,280) | 0.71 (0.59, 0.85)* | 3.0% | (132/4,402) | 0.72 (0.59, 0.87)* |
|  | 60 and over | 2.8% | (62/2,233) | 0.62 (0.48, 0.81)* | 2.7% | (65/2,402) | 0.66 (0.51, 0.86)* |
|  | Overall | 4.2% | (778/18,531) | - | 3.9% | (759/19,594) | - |
| Overall (Both VCH and IH combined) | Urban | 4.6% | (1,290/28,102) | 1.44 (1.29, 1.60)* | 4.2% | (1,207/29,013) | 1.67 (1.48, 1.88)* |
|  | Rural | 3.2% | (470/14,547) | 0.69 (0.62, 0.77)* | 2.5% | (372/14,660) | 0.60 (0.53, 0.67)* |
|  | LPN / Care Aides | 5.8% | (684/11,853) | 1.69 (1.53, 1.87)* | 5.0% | (609/12,066) | 1.68 (1.51, 1.86)* |
|  | Nurses | 3.8% | (483/12,788) | 0.88 (0.79, 0.98)* | 3.2% | (427/13,212) | 0.85 (0.76, 0.95)* |
|  | Administration | 3.2% | (232/7,179) | 0.74 (0.64, 0.85)* | 3.0% | (216/7,246) | 0.79 (0.68, 0.91)* |
|  | Allied Health | 2.8% | (179/6,289) | 0.64 (0.55, 0.75)* | 2.5% | (162/6,400) | 0.66 (0.56, 0.77)* |
|  | Support | 4.1% | (158/3,873) | 0.99 (0.84, 1.17) | 3.5% | (134/3,866) | 0.95 (0.80, 1.14) |
|  | 39 and under | 5.0% | (909/18,235) | 1.45 (1.32, 1.60)* | 4.4% | (832/18,762) | 1.50 (1.36, 1.66)* |
|  | 40-49 | 4.0% | (399/9,854) | 0.97 (0.87, 1.09) | 3.3% | (331/9,954) | 0.89 (0.79, 1.01) |
|  | 50-59 | 3.5% | (335/9,633) | 0.80 (0.71, 0.90)* | 3.1% | (301/9,787) | 0.81 (0.71, 0.92)* |
|  | 60 and over | 2.4% | (117/4,927) | 0.53 (0.44, 0.65)* | 2.2% | (115/5,170) | 0.58 (0.47, 0.70)* |
|  | Overall | 4.1% | (1,760/42,649) | - | 3.6% | (1,579/43,673) | - |

\* = unadjusted odds ratio (OR) is significant at 95% confidence

It could be seen that a larger proportion of HCWs living in urban settings were vaccinated compared to their rural counterparts overall (Table 1); however, they also had a cumulatively higher rate of PCR-confirmed SARS-CoV-2 (Table 2). Table 1 further shows a higher rate of unvaccinated rural-dwelling workers (11.1% urban vs 13.2% rural; odds ratio 0.82; 95% CI 0.76, 0.89; p < 0.001). Those dwelling rurally and employed by VCH were more than twice as likely to be unvaccinated both on September 12, 2021, the day before the mandate was announced for the entire healthcare workforce (odds ratio 2.25; 95% CI 1.85, 2.74; p < 0.001) and October 27, 2021, when *this* mandate came into effect (odds ratio 2.89; 95% CI 2.20, 3.79; p < 0.001). A separate analysis conducted of only the subset of healthcare workers who work in long-term care (LTC) facilities, using August 12^th^, the date of the announcement that all *LTC* workers would require vaccination, the rate of first doses was indeed shown to significantly increase, but 177 of 5736 (3.1%) LTC workers remained unvaccinated at the time the mandate came into effect, and importantly, 116 (66.5%) of unvaccinated LTC workers were in rural areas.

Before the mandate announcement (September 12, 2021), the SARS-CoV-2 infection rate was significantly lower for IH than VCH (3.4% IH versus 3.9% in VCH; odds ratio 0.87; 95% CI 0.79, 0.97; p = 0.009); rural workers indeed had a significantly lower infection rate across both health authorities (2.5% versus 4.2% among urban counterparts; odds ratio 0.60; 95% CI 0.53, 0.67; p < 0.001). Worrisomely, a full 12.2% of HCWs in Interior Health were unvaccinated compared to 3.6% in VCH (odds ratio 3.76; 95% CI 3.46, 4.09; p < 0.001), despite all being subject to the same provincial policies. The relative difference between health authorities continued to October 27, 2021, when 6.5% of HCWs in Interior Health were still unvaccinated compared to only 1.6% in VCH (odds ratio 4.17; 95% CI 3.68, 4.72; p < 0.001).

Regarding question #3, Table 1 also shows these indicators by occupational role. It can be seen that 5.8% of LPN/care aides in IH had contracted PCR-confirmed SARS-COV-2 infections, compared to 4.1% of the IH healthcare workforce overall. In VCH, the corresponding figures were 5.7% and 4.2%. Within IH across both time points, LPN/care aides, administrative and support workers had significantly lower vaccination rates, and nurses and allied health workers had higher vaccination rates. Within VCH, only support workers had a significantly lower vaccination rate, and nurses with a higher vaccination rate. With respect to risk of SARS-CoV-2 infections we see that only LPNs/care aides have higher infection rates, in both health authorities, at both time points (Table 2).

For question #4, considering the differences between age groups, within IH, the vaccinated rate in HCWs was significantly lower in the age group 30-39 years across both time points; simultaneously, the SARS-CoV-2 infection rate was significantly higher in that age group at both time points. in VCH, differences in vaccination by age group did not appear consistent between time points; however, when we considered SARS-CoV-2 infection rates, we found that the infection rate was higher for those aged 39 and under (Table 2). Analysing the entire workforce of both regions combined to determine differences between urban and rural dwelling workers, taking age and occupational mix into consideration, we found that rural workers were vaccinated at a significantly lower rate than their age-adjusted, occupation-adjusted counterparts in urban areas, both by September 12^th^ before the mandate was announced (odds ratio 0.53; 95% CI 0.49, 0.57; p < 0.001), and by October 27^th^ when the mandate came into force (odds ratio 0.54; 95% CI 0.49, 0.59; p < 0.001).

Regarding question #5 (the extent to which infection rates drove vaccine uptake in HCWs), considering the average community infection rate over the 2 weeks prior to receiving the first dose using conditional logistic regression, we found that an average increase of 1 case per 100,000 in the community SARS-CoV-2 infection rate was associated with a 3.5% (95% CI 3.2, 3.8%) increased likelihood of vaccination two weeks later.

Regarding question #6, the extent to which the announcement of the provincial vaccine mandate requiring all BC healthcare workers to be vaccinated before October 27, 2021 drove up vaccination rates, ITS segmented regression analysis of the period from July 1, 2021 to October 27 showed significant effects over the vaccine mandate period, with similar effects in both urban and rural settings (Table 3). However, while the daily proportion of unvaccinated workers who received first doses showed an *immediate* rate increase of 0.78%, (from 1.01% vaccinating per day to 1.79%), the *sustained* effect was a daily reduction of 0.028% HCWs being vaccinated each day after the announcement. This showed a sustained cumulative effect of -1.29% over the 45-day period between the mandate announcement and implementation, such that the overall impact of the mandate on first dose uptake was unclear (Figure 4). When second doses were considered, the immediate effect was not significant, but the sustained effect showed a significant increase, as would be expected (Figure 5). The sustained effect rate increase of 0.063% second doses daily after the mandate announcement showed a sustained cumulative effect of 2.77% over the period.

**Table 3:** Effect of the vaccine mandate on the vaccination rate for both first and second doses, using segmented regression ITS analysis

| Subgroup | First doses |  | Second doses |  |
| --- | --- | --- | --- | --- |
|  | Immediate effect<br>(95% CI) | Sustained effect<br>(95% CI) | Immediate effect<br>(95% CI) | Sustained effect<br>(95% CI) |
| IH | 0.67% (0.22, 1.11)* | -0.028% (-0.044, -0.013)* | 0.15% (-0.23, 0.53) | 0.055% (0.042, 0.068)* |
| VCH | 1.25% (0.48, 2.01)* | -0.023% (-0.050, 0.003) | -0.19% (-0.71, 0.33) | 0.094% (0.076, 0.112)* |
| Urban | 0.68% (0.20, 1.17)* | -0.021% (-0.037, -0.004)* | 0.05% (-0.33, 0.44) | 0.064% (0.051, 0.077)* |
| Rural | 0.91% (0.38, 1.43)* | -0.037% (-0.055, -0.019)* | 0.13% (-0.33, 0.58) | 0.061% (0.045, 0.076)* |
| LPN / Care Aides | 0.82% (0.22, 1.42)* | -0.027% (-0.047, -0.006)* | -0.14% (-0.53, 0.24) | 0.052% (0.039, 0.065)* |
| Nurses | 0.64% (0.15, 1.13)* | -0.030% (-0.047, -0.013)* | 0.24% (-0.18, 0.65) | 0.045% (0.031, 0.060)* |
| Administration | 0.65% (0.12, 1.18)* | -0.013% (-0.031, 0.006) | 0.33% (-0.24, 0.91) | 0.087% (0.067, 0.107)* |
| Allied Health | 1.05% (0.48, 1.61)* | -0.040% (-0.059, -0.021)* | -0.10% (-0.68, 0.48) | 0.065% (0.045, 0.085)* |
| Support | 1.18% (0.45, 1.92)* | -0.046% (-0.072, -0.021)* | -0.06% (-0.62, 0.50) | 0.065% (0.046, 0.084)* |
| 39 and under | 0.68% (0.16, 1.20)* | -0.033% (-0.051, -0.015)* | 0.05% (-0.33, 0.44) | 0.060% (0.046, 0.073)* |
| 40-49 | 0.73% (0.14, 1.31)* | -0.022% (-0.043, -0.002)* | 0.11% (-0.39, 0.60) | 0.065% (0.048, 0.082)* |
| 50-59 | 1.02% (0.39, 1.64)* | -0.028% (-0.050, -0.007)* | 0.16% (-0.34, 0.67) | 0.072% (0.055, 0.089)* |
| 60 and over | 1.01% (0.49, 1.52)* | -0.016% (-0.034, 0.001) | 0.19% (-0.34, 0.73) | 0.057% (0.039, 0.075)* |
| **Overall** | **0.78% (0.31, 1.25)\*** | **-0.028% (-0.044, -0.012)\*** | **0.09% (-0.29, 0.47)** | **0.063% (0.050, 0.076)\*** |

\* = effect of the mandate compared is significantly different from 0 at 95% confidence. Values in red are negative. No subgroup is significantly different from the other subgroups.

**Figure 4:**
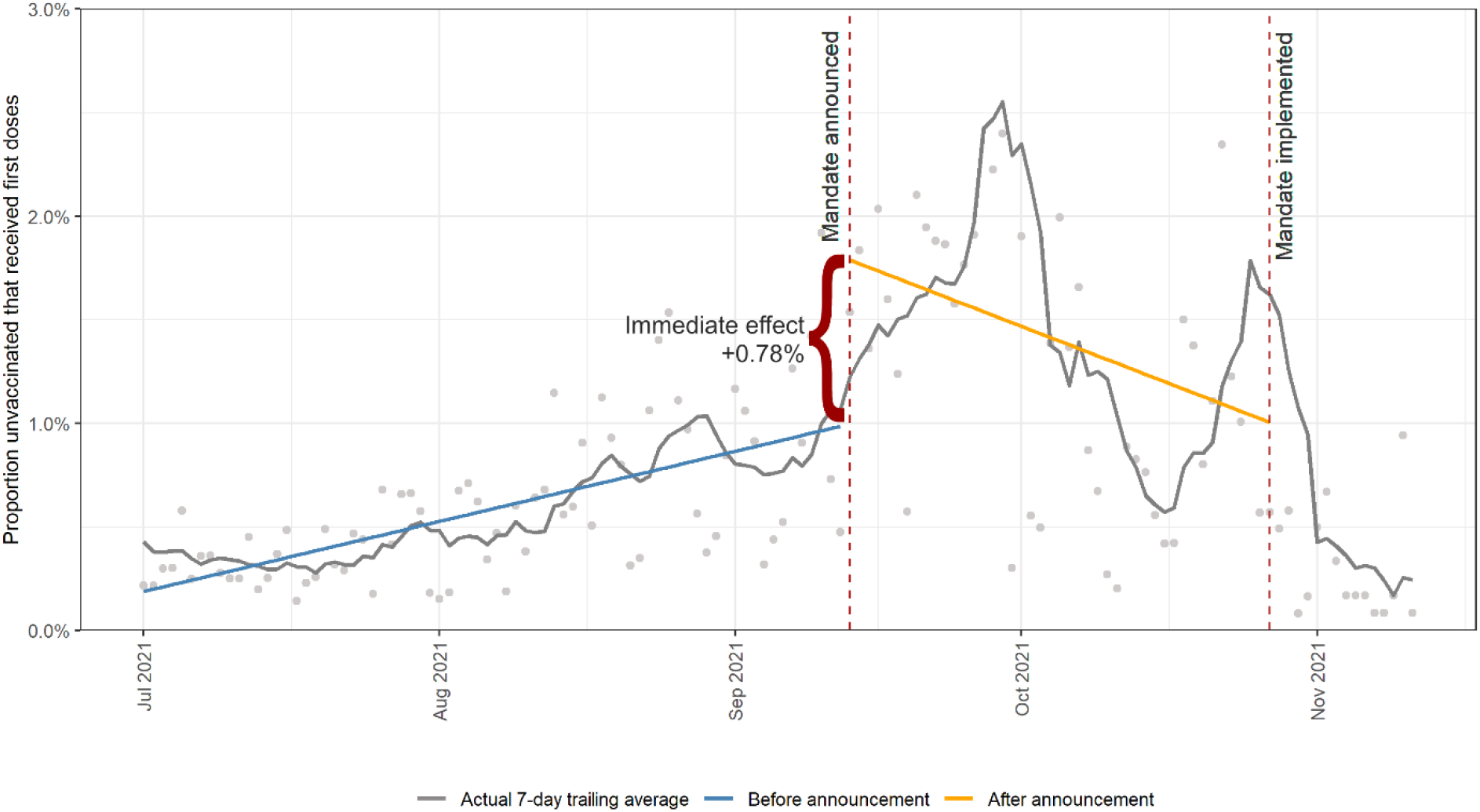
Overall daily proportion of unvaccinated who received <u>first doses</u> from July 1, 2021, to November 11, 2021, with the segmented regression ITS predicted values (blue and orange lines). The immediate effect of 0.78% is the difference between the two segments on the date of the announcement.

**Figure 5:**
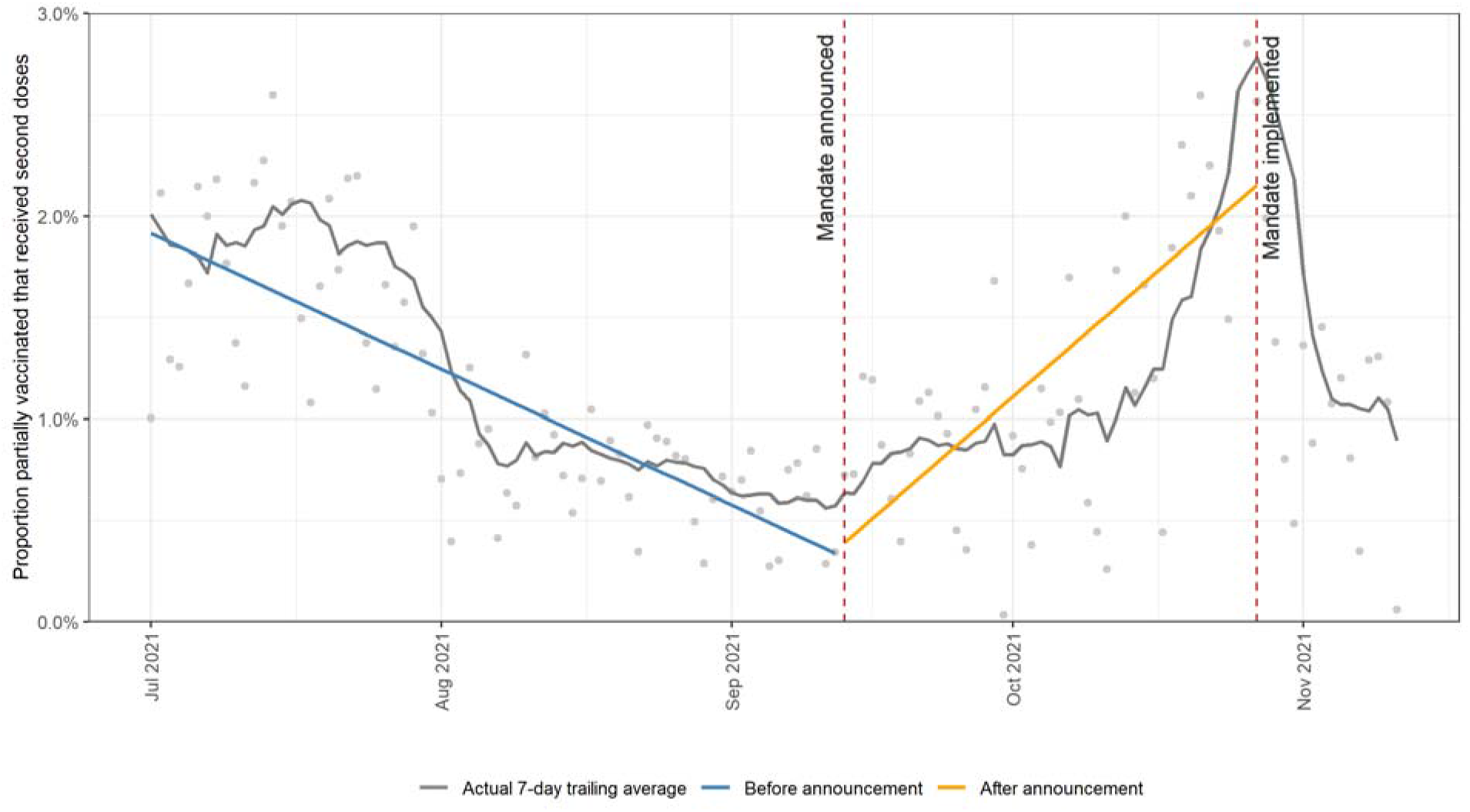
Overall daily proportion of partially vaccinated workers who received <u>second doses</u> from July 1, 2021, to November 11, 2021, with the segmented regression ITS predicted values.

## Discussion

We found, as have others^14^, that vaccine uptake was significantly lower in those who live in rural areas compared to their urban counterparts. There may be more hurdles to being vaccinated in a rural location than in a more populated region including having to drive to a vaccine appointment, perhaps in a neighbouring town, which could deter some people from being vaccinated. Nonetheless, the rural-urban divide in vaccine uptake by HCWs is concerning. Globally, HCWs have faced a heavy emotional and physical toll during the pandemic. We found that COVID-19 rates were higher in some occupational groups of HCWs than others, as has also been reported elsewhere^28^. This has resulted in those working on the frontlines of healthcare to quit in unprecedented numbers^30-32^. We found an association between vaccine uptake by HCWs and HCW COVID-19 rates in the preceding period as would be expected, although the higher rates of COVID-19 infection in some occupational groups did not lead to increased vaccination in these groups.

For some HCWs who may be vaccine hesitant, mandates have exacerbated an already stressful situation^33^. In some jurisdictions, vaccine mandates have been highly effective in driving up vaccinations; in France, the law on mandatory vaccination for HCWs led to a massive boost in vaccination rates, from 60% in July (when the new requirement was announced) to over 99% in October^34^. France, in mandating vaccination for most professions, moved from a country with a high case count and low vaccination rates to one of the countries with the highest vaccination rates in the world: France’s vaccination rates markedly increased, and its COVID-19 cases accordingly declined ^35^. However, it is important to note the other side of the coin to vaccine mandates for HCWs^36^ – whilst vaccination has increased, those who chose to not be vaccinated lost their jobs and were lost to the healthcare system^37^. The effects of this have been felt more in small towns and rural locations^37,38^ which potentially were already suffering from staff-shortages. In our study, over 1,800 workers, comprising 6.4% of rural HCWs and 3.3% of urban HCWs, remained unvaccinated despite consequent employment termination. This loss of resources during an already stressed time could have devastating effects on the provision of healthcare – particularly in rural communities.

In March 2021, a poll by the Kaiser Family Foundation in the US found that 21% of rural residents stated that they would “definitely not” get a vaccine compared with 10% of urban residents. Among the rural respondents, 45% of younger adults (aged 18–64 years) stated that they would “definitely not” get a vaccine compared with 8% of older adults (aged 60–69 years)^39^. Our data from British Columbia, Canada, echoed the greater vaccine hesitancy in rural compared to urban areas. Rural, remote and Northern communities in Canada have a unique risk profile for COVID-19; and while geographic barriers in remote areas can provide some protection from exposure if visitors are prevented from entering the community, the virus is spread more easily due to poverty, crowded housing, and lack of clean and running water for sanitation, which are conditions on many First Nations communities^40,41^. Many rural communities have poor communication technology, which limits the spread of information about disease prevention, and can lead to an increase in stigma and inappropriate response to the disease, as well as decreased ability of health professionals to connect with cases and their contacts^42^. Importantly, contrary to concern that spreading the disease to others would be greater in more densely populated areas thereby driving vaccination rates, Jung and Albarracin^43^ showed the opposite to be true in their study, whereby the less-densely populated areas were more motivated to get vaccinated based on social concerns than their urban counterparts.

A US national survey found that the people who believed the COVID-19 vaccine was unsafe, were less willing to receive the vaccine, knew less about the virus and were more likely to believe COVID-19 vaccine myths, on average were less educated, had lower income, and were more predominantly rural dwelliers^44^. Other studies have noted that the vast majority of rural residents reported that they trust their own healthcare providers for information on COVID-19 vaccines, which highlights the importance of public health practitioners working with established and locally trusted providers in outpatient health care systems in rural areas^45^. Similar to other studies^46^ we expected mandates to drive up vaccination rates. In a recently published study of 6 countries, it was noted that countries with pre-intervention vaccine uptake below average had a more pronounced increase in daily vaccinations following mandatory COVID-19 certificates compared with those where uptake was already average or higher^36^. As such, it would have been expected that the BC mandate for HCWs would have significantly narrowed the gap in vaccine uptake between rural and urban HCWs. We found, however, that making vaccination mandatory to work in healthcare may indeed have increased vaccine rates in HCWs in BC, but fell short of achieving very high levels of uptake. A different research design would be needed to ascertain reasons for non-vaccination, as well as the specific impact of the announcement of the LTC mandate on the healthcare workforce as a whole. Our results nonetheless suggest that the vaccine mandate had some impact among those hesitant to be vaccinated albeit not on those who decisively rejected vaccination, and did have a significant, albeit small, effect on uptake of second doses. In any case, given that rural areas had a lower vaccine uptake prior to the mandate, the fact that more rural HCWs still remained unvaccinated after the mandate is worrisome.

Given the ongoing pandemic, and now even greater concern and uncertainly regarding new emerging variants, such as Omicron^8^, further research is needed to better understand the reasons behind ongoing vaccine hesitancy and what can be done to address these factors. The analysis presented here was conducted based on data ending just before Omicron spread rapidly in this jurisdiction; further analysis is now needed to assess the impact on vaccine uptake given the lower effectiveness of the vaccine against Omicron^47^ and possible requirements for boosters. Specifically It is crucial that we acquire a deep understanding of how rurality impacts the 7C’s of vaccine hesitancy ^48^ (*complacency*: not perceiving diseases as high risk enough to bother taking action; *constraints*: structural and psychological barriers; *confidence*: trust in the effectiveness and safety of vaccines, the system that delivers these and/or motivations of policymakers; *calculation*: calculating one’s own risk; and aspects pertaining to *collective responsibility, i*.*e*. willingness to protect others; as well as *conspiracy*: the tendency to endorse conspiratorial beliefs about vaccination; and *compliance:* the tendency to adhere to regulations.) Moreover, with ongoing boosters possibly essential to protect the health of the public, it is necessary that attention be paid to how to increase uptake of vaccinations in rural healthcare workers without aggravating staff shortages in these areas. Given that rural HCWs’ beliefs, behaviours and actions are reflective of their communities, there may be value in examining the impacts of rural community-based strategies at the local level with the view to improving the effectiveness of vaccination uptake and other public health/health literacy initiatives/campaigns. Intervention studies exploring the use of trusted local leaders and the impact on vaccine uptake are urgently needed.

## Data Availability

All data produced in the present study are available upon reasonable request to the authors.

## Acknowledgments

The authors wish to offer our gratitude to all the healthcare workers who have worked tirelessly throughout the pandemic. We also thank the many University of British Columbia medical students who staffed the Physician Occupational Safety and Health (POSH) service, as well as the Infection Prevention and Control, Public Health and People Safety teams who all work tirelessly to ensure that the healthcare workforce was well-protected. We also would like to thank the teams who extracted the data we needed for this research.

---

**Table S1:** Mapping of local health areas (LHAs) to their urban / rural categorization. 480 and 265 workers from IH and VCH respectively had no local home address listed in the database.

| Health Authority | Type | LHA (population) | HCW home population |
| --- | --- | --- | --- |
| Interior Health (IH) | Urban | Central Okanagan (219,454); Kamloops (129,286) | 11,643 |
|  | Rural | Vernon (74,177); Penticton (46,248); Salmon Arm (37,935); Nelson (28,128); Cranbrook (27,982); Cariboo/Chilcotin (26,754); Trail (21,991); Southern Okanagan (21,897); Fernie (18,178); 100 Mile House (16,183); Castlegar (15,074); Creston (13,557); Merritt (13,370); Summerland (13,319); Armstrong/Spallumcheen (11,329); Windermere (11,135); Revelstoke (10,645); Kimberley (10,356); Grand Forks (10,019); Enderby (8,212); Golden (8,092); South Cariboo (7,233); Princeton (6,080); Keremeos (5,743); Lillooet (5,278); Arrow Lakes (5,152); North Thompson (4,317); Kettle Valley (3,833); Kootenay Lake (3,818) | 13,146 |
| Vancouver Coastal Health (VCH) | Urban | Abbotsford; Burnaby; Chilliwack; Delta; Langley; Maple Ridge/Pitt Meadows; Mission; New Westminster; North Vancouver; Richmond; South Surrey/White Rock; Surrey; Tri-Cities; Vancouver; West Vancouver/Bowen Island | 18,328 |
|  | Rural | Agassiz/Harrison; Central Coast; Hope; Howe Sound; Powell River; Sunshine Coast | 1,937 |

**Table S2:** Weightings for calculating the age and region-adjusted community infection rate on November 11, 2021.

| Worksite health authority | Residence health authority | Age group | HCW: age and region proportion (p<sub>1</sub>) | Public: age and region proportion (p<sub>2</sub>) | Weighting (p<sub>1</sub> / p<sub>2</sub>) |
| --- | --- | --- | --- | --- | --- |
| Interior | Interior | <10 | 0.0% | 9.3% | 0.000 |
|  |  | 10-19 | 0.6% | 10.0% | 0.059 |
|  |  | 20-29 | 16.7% | 10.3% | 1.626 |
|  |  | 30-39 | 26.6% | 11.1% | 2.388 |
|  |  | 40-49 | 22.6% | 11.7% | 1.931 |
|  |  | 50-59 | 22.2% | 16.2% | 1.365 |
|  |  | 60-69 | 10.2% | 15.9% | 0.641 |
|  |  | 70-79 | 1.1% | 9.6% | 0.120 |
|  |  | 80-89 | 0.0% | 4.7% | 0.000 |
|  |  | 90+ | 0.0% | 1.0% | 0.000 |
| VCH | Fraser | <10 | 0.0% | 6.6% | 0.000 |
|  |  | 10-19 | 0.1% | 7.1% | 0.008 |
|  |  | 20-29 | 5.9% | 7.8% | 0.750 |
|  |  | 30-39 | 8.5% | 8.1% | 1.049 |
|  |  | 40-49 | 7.0% | 8.5% | 0.833 |
|  |  | 50-59 | 6.9% | 9.0% | 0.763 |
|  |  | 60-69 | 2.9% | 7.0% | 0.415 |
|  |  | 70-79 | 0.2% | 3.9% | 0.059 |
|  |  | 80-89 | 0.0% | 1.9% | 0.003 |
|  |  | 90+ | 0.0% | 0.5% | 0.000 |
|  | VCH | <10 | 0.0% | 3.3% | 0.000 |
|  |  | 10-19 | 0.2% | 3.7% | 0.045 |
|  |  | 20-29 | 10.7% | 5.9% | 1.798 |
|  |  | 30-39 | 18.0% | 6.0% | 3.012 |
|  |  | 40-49 | 15.6% | 5.7% | 2.755 |
|  |  | 50-59 | 15.3% | 5.9% | 2.589 |
|  |  | 60-69 | 8.0% | 4.8% | 1.675 |
|  |  | 70-79 | 0.9% | 2.6% | 0.351 |
|  |  | 80-89 | 0.0% | 1.4% | 0.017 |
|  |  | 90+ | 0.0% | 0.4% | 0.000 |

Proportions are calculated within worksite health authority.

